# People living with multiple long-term conditions have different pathways of unscheduled care in hospital: findings from an analysis of routinely-collected clinical data

**DOI:** 10.64898/2026.08.28.26361696

**Authors:** Miles D Witham, Felicity Evison, Sue Bellass, Rachel Cooper, Suzy Gallier, Sara Pretorius, Elizabeth Sapey, Jana Suklan, Avan A Sayer, ADMISSION Research Collaborative

## Abstract

**Study Objective:** Little is known about where in hospital care for multiple long-term conditions (MLTC) is delivered. We aimed to describe pathways of care (ward transfers) and outcomes for people admitted to hospital for unscheduled care by MLTC status and other key sociodemographic characteristics.

**Design and setting:** Analysis of routinely-collected electronic health records from a large acute UK hospital.

**Participants:** Adult unscheduled care admissions from 1st July 2018 to 30th June 2019. The presence of two or more of 59 long-term conditions was ascertained using ICD-10 codes from previous hospital discharges.

**Main outcome measures:** Markov state transition probabilities were derived for ward moves and compared for MLTC vs no MLTC, age, sex, ethnicity and neighbourhood deprivation. Outcomes (length of stay, death, readmission, move from definitive ward) and time spent in emergency and assessment departments were compared between subgroups.

**Results:** A total of 33,252 adults, mean age 56.0 (SD 21.9) years were analysed; 14,834 (42.4%) had MLTC. People with MLTC were more likely to die in hospital (4.2 vs 1.9%, p<0.001), transfer to internal medicine wards or older people’s medicine wards, were less likely to transfer to surgical wards, had longer median length of stay (1.83 vs 0.69 days, p<0.001), stayed longer in acute medical units (15.5 vs 9.6 hours, p<0.001), and were more likely to move from their definitive ward (18.2 vs 16.4%, p=0.002).

**Conclusion:** Unscheduled hospital care pathways are complex and differ for people with MLTC, who have worse outcomes and may be less likely to receive optimal care.

## Introduction

Multiple long-term conditions (MLTC, also referred to as multimorbidity) are a major challenge facing 21st century medicine [1] and are an important cause of admission to hospital [2]. People with MLTC experience longer length of hospital stay, worse outcomes and lower satisfaction with care than people with single conditions admitted to hospital [3–5]. Whilst there is agreement that current hospital-based systems are not fit for purpose in delivering person-centred, coordinated care for people living with MLTC [6], empirical data describing pathways of care for people with MLTC admitted to hospital are lacking. Understanding where people receive care in hospitals and how this might differ for people with and without MLTC is an essential prerequisite for governments and healthcare providers to undertake effective redesign of healthcare systems and care pathways.

The ADMISSION Research Collaborative is a UK-based programme set up specifically to research MLTC in people admitted to hospital [7]. Understanding the impact of MLTC on pathways of unscheduled care is of particular importance: unscheduled care admissions drive much of the current overloading of the UK hospital system [8]; people living with MLTC are more likely to require unscheduled care [9] and unscheduled care admissions are associated with higher mortality rates and longer length of stay than elective admissions [10].

Analysis of routinely-collected clinical data (via pathway mapping and process mining) has been used previously to understand the sequence of care actions during either single episodes of care or across the course of an illness [11]. We have previously shown the feasibility of using routinely-collected clinical data to map ward transfers for people admitted with an exemplar condition: exacerbation of COPD [12]. The aim of this work was to describe pathways of care (as depicted by ward transfers) and outcomes for people experiencing unscheduled admissions to hospital by MLTC status and other key sociodemographic characteristics.

## Methods

### Design and setting

We conducted an analysis of routinely-collected hospital electronic health records held by the Health Data Research (HDR) UK Acute Care Hub (PIONEER) [13]. We studied data from unscheduled care admissions to the Queen Elizabeth Hospital Birmingham (QEHB), an acute hospital admitting adults and serving a large urban area in the West Midlands, UK. QEHB has 1269 beds including 80 level 2/3 intensive care unit (ICU) beds, an emergency department that assesses >300 people per day (with 100 per day admitted for further assessment), and a mixed secondary and tertiary practice that includes all major adult specialties except for obstetrics and gynaecology. Electronic health records (EHR) at QEHB are held by a bespoke system (PICS, Birmingham Systems, Birmingham, UK) and contain time-stamped, structured data including demography, location, time of admission and discharge, all treatments and investigations, and physiological measures. The EHR has been in place since 1999.

### Data source

For this analysis, we first identified data for all unscheduled care admissions to QEHB from 1^st^ July 2018 to 30^th^ June 2019. We chose these dates as the most recent dates where the COVID-19 pandemic would not have disrupted patterns of care and would not have interacted with the one-year follow up period; we chose a time period encompassing a complete winter period. We confined the study population to people who were admitted for unscheduled care (i.e. excluding all those admitted for elective or planned care, including renal dialysis, chemotherapy and radiotherapy). We further confined the analysis to people presenting to the emergency department, acute medical assessment suite or surgical wards, excluding those whose first point of attendance was outwith this pathway, both to ensure included people were definitely admitted for unscheduled care and to reduce the number of initial pathways with small numbers to analyse. People attending the emergency department but who were discharged home without overnight admission were not included in this analysis. We identified the first unscheduled care admission for each individual during the study period; this admission was used as the index episode for analysis. For outcomes (hospital readmission, death), we extracted data from the EHR for a one-year period starting from the date of admission for the index episode.

### Derivation of key variables

To ascertain diagnoses, we used a previously developed list of 60 long-term conditions for this analysis [14]. The ADMISSION 60-condition list was based on the results of a previous Delphi consensus exercise [15], with subsequent amendments identified in collaboration with the ADMISSION Patient Advisory Group to optimise the list for use in hospital-based MLTC research. ICD-10 codes published as part of this 60-condition list were used to extract and operationalise diagnoses recorded in hospital discharge diagnosis lists prior to the index admission, with the exception of recurrent urinary tract infection which required at least two admissions with a relevant code within a six-month time period. The presence of MLTC was operationalised as two or more long-term conditions from this list (with the exception of HIV, for which data are redacted by PIONEER), in line with current consensus statements [16], thus data on 59 conditions were included in these analyses. No limitation was placed on minimum duration living with long-term conditions for these analyses.

Moves between wards were identified from time-stamped data held within the EHR for the index admission. Ward durations of less than 30 minutes were considered unlikely to be true moves and were attributed to clerical error. Where overlaps of wards occurred, a review of the observations and medication administrations during that time was undertaken and the ward location from those records deemed to be the true location. An ontology of ward types was created in collaboration with clinical colleagues to collapse the several hundred places of care down to a tractable series of categories of places of care likely to be of clinical relevance (Supplementary Table 1). Duration of stay in each place of care was calculated (in hours) and the total summed to provide the overall length of stay for the index admission (in days). We excluded transfers to intervention suites (e.g. endoscopy, interventional radiology) and temporary transfers for renal dialysis whilst an inpatient. To interrogate the phenomenon of ‘boarding’ (a move from one ward to another for reasons of capacity management rather than for compelling clinical need) [17], the ‘definitive ward’ was defined as the first ward that an individual was moved to that was not the emergency department, acute medical unit or critical care unit.

Death during the one-year follow-up period was extracted from the EHR and linked national spine record for out-of-hospital deaths. Where death occurred during the index hospital stay this was ascertained via the discharge record. Sociodemographic data were extracted from the EHR, using age and sex (as recorded at birth) held at the time of the index admission. Deprivation was recorded as fifths of the Index of Multiple Deprivation (IMD) [18] derived from partial postcodes, using the postcode recorded at the time of index admission. Ethnicity was extracted from codes in the EHR denoting 18 standard categories recommended by the UK National Health Service [19]. Individuals missing ethnicity, and missing IMD status (e.g. people without homes, those in the armed forces who did not have postcodes) are recorded as ‘not known’ in demographics tables; these categories have been excluded from the comparative analyses as they are likely not to be missing at random.

### Analysis

Transitions between different categories of wards were envisaged as states in a Markov transition matrix. The probability of transition between each category of ward or care setting and each other category (including the probability of transition to death and to discharge) were calculated and depicted in matrix tables as the number of transitions was too large to easily depict graphically on state transition diagrams. For this analysis, the length of time spent in a particular ward category or care setting was not included in the analysis. Groups with small numbers (n<5) were suppressed in line with PIONEER governance stipulations [13]. Continuous variables were compared using Student’s t-test (for normally distributed continuous variables) or Mann-Whitney U test (for non-normally distributed continuous variables). Categorical variables were compared using Pearson’s chi-squared test. A two-sided p value of <0.05 was taken to indicate statistical significance for all analyses. All analyses were conducted using R Statistical Software (v4.1.2; R Core Team 2021, Vienna, Austria).

The study was conducted under ethical approval provided for PIONEER, the Health Data Research Hub for Acute Care (East Midlands–Derby [20/EM/0158] and Confidentiality Advisory Group (20/CAG/0084). People who had chosen to opt out of data use for research and planning via the NHS National Data Opt-Out or locally were excluded. Results were presented to the ADMISSION Patient Advisory Group for their feedback and interpretation, with input into the paper provided by ADMISSION public coapplicants (VB and RH).

## Results

A total of 33,252 individuals were admitted at least once for unscheduled care in the analysis period and were included in analyses (Figure 1). Descriptive data on the sample are shown in Supplementary Table 2, and data on the prevalence of recorded long-term conditions are given in Supplementary Table 3. The prevalence of MLTC was high (14,834; 42.4%).

**Figure 1.**
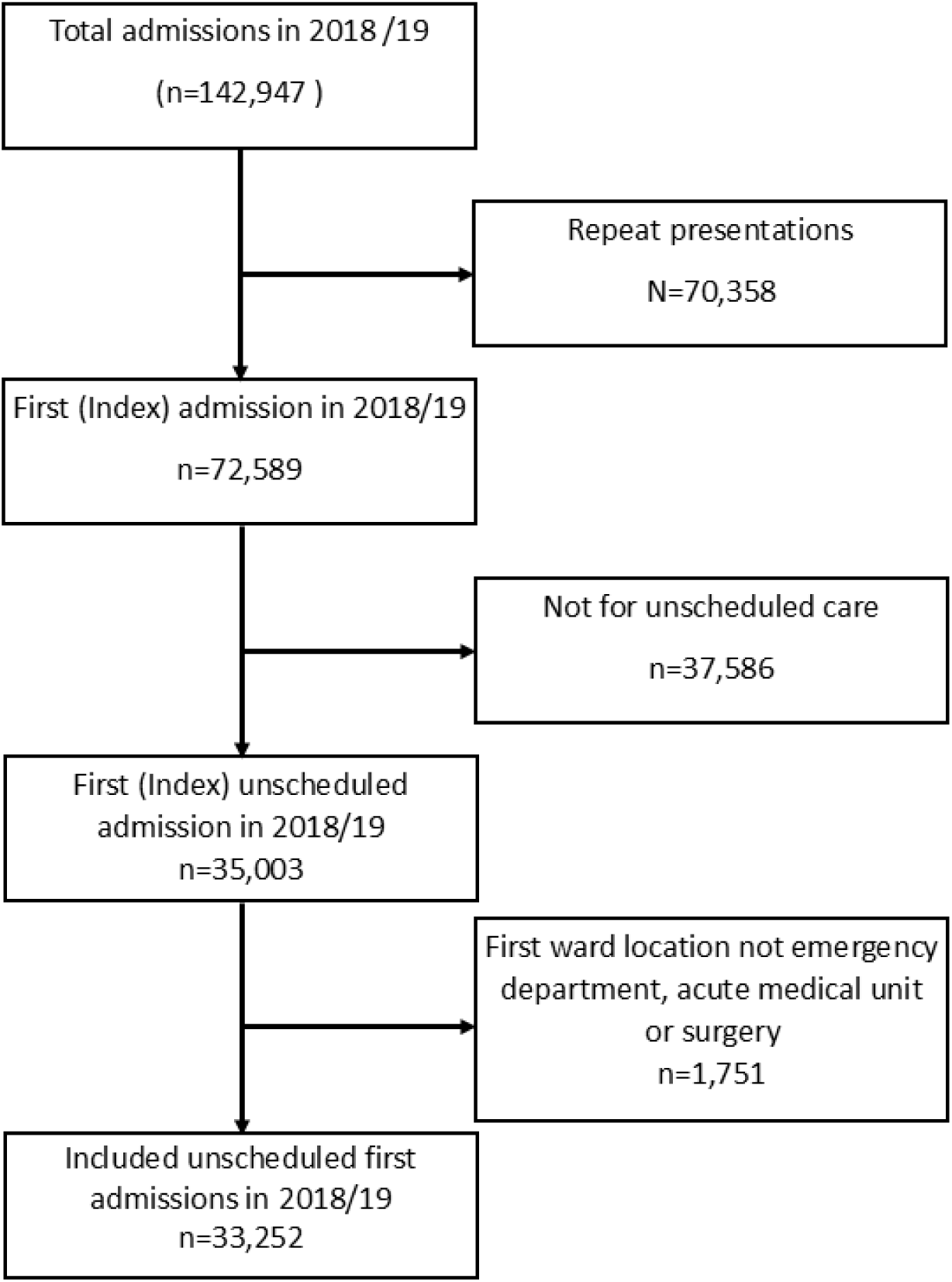
Flowchart of individuals included in analysis.

Key outcomes associated with hospital stay are shown in Table 1. MLTC was associated with longer length of stay in hospital, higher in-hospital and one-year mortality, and higher rates of 30-day readmission to hospital, but no difference in the median number of places of care was evident between those with and without MLTC. Similar associations were seen between age and these outcomes. Greater number of long-term conditions was associated with older age as expected (Supplementary Figure 1). Men were more likely than women to be admitted to the intensive care unit but no difference in the number of places of care was evident between sexes.

**Table 1.** Relationship between MLTC and other individual descriptors, and key outcomes (number of wards, critical care admission, length of stay, death and readmission)

|  | Median number of wards<br>during admission (IQR) | Admitted to ICU during<br>admission (n, %) | Median LOS (days)<br>(IQR) | Death during hospital<br>admission (n, %) | Death within a year of<br>admission (n, %) | Readmission to hospital<br>within 30 days of discharge |
| --- | --- | --- | --- | --- | --- | --- |
| All individuals | 2 (2, 3) | 982 (3.0%) | 1 (0.29, 4.63) | 951 (2.9%) | 3459 (10.4%) | 4749 (14.3%) |
| *Presence of MLTC* |  |  |  |  |  |  |
| No MLTC (0-1 conditions) | 2 (2, 2) | 641 (3.3%) | 0.69 (0.25, 3.04) | 360 (1.9%) | 1031 (5.4%) | 2276 (11.8%) |
| MLTC (2 or more conditions) | 2 (2, 3) | 341 (2.4%) | 1.83 (0.38, 7.00) | 591 (4.2%) | 2428 (17.3%) | 2473 (17.6%) |
| *Age (years)* |  |  |  |  |  |  |
| <=70 | 2 (2, 2) | 714 (3.1%) | 0.67 (0.25, 2.92) | 248 (1.1%) | 928 (4.0%) | 3046 (13.3%) |
| >70 | 2 (2, 3) | 268 (2.6%) | 2.88 (0.63, 9.88) | 703 (6.8%) | 2531 (24.5%) | 1703 (16.5%) |
| *Sex* |  |  |  |  |  |  |
| Men | 2 (2, 3) | 658 (4.1%) | 1.08 (0.29, 4.83) | 474 (3.0%) | 1706 (10.7%) | 2328 (14.5%) |
| Women | 2 (2, 3) | 324 (1.9%) | 0.92 (0.29, 4.29) | 477 (2.8%) | 1752 (10.2%) | 2421 (14.0%) |
| *Fifth of deprivation* |  |  |  |  |  |  |
| 1 (most deprived) | 2 (2, 3) | 443 (2.7%) | 0.96 (0.29, 4.17) | 411 (2.6%) | 1528 (9.5%) | 2396 (14.9%) |
| 2 to 5 | 2 (2, 3) | 511 (3.0%) | 1.04 (0.29, 4.96) | 536 (3.2%) | 1920 (11.4%) | 2287 (13.6%) |
| *Ethnicity* |  |  |  |  |  |  |
| White | 2 (2, 3) | 670 (3.2%) | 1.13 (0.29, 5.42) | 617 (2.9%) | 2388 (11.2%) | 3221 (15.2%) |
| Non-white | 2 (2, 2) | 169 (2.1%) | 0.67 (0.25, 3.08) | 101 (1.3%) | 438 (5.5%) | 1105 (13.9%) |

Supplementary Figure 2 shows pathways followed by people moving between different wards in hospital, and Table 2 summarises the transition probabilities of moving from one place of care to another, or to discharge, or to death, during the hospital stay for the whole cohort. The majority of people seeking unscheduled care entered the hospital system via the emergency department, and the majority of these transferred to the acute medical unit as their next place of care.

**Table 2.** Summary of the probability of transition between different wards for the whole analysis cohort.

|  |  | *Transition end point* |  |  |  |  |  |  |  |  |
| --- | --- | --- | --- | --- | --- | --- | --- | --- | --- | --- |
|  |  | ED | AMU | Surgical | Speciality Med | Internal Med | Older Peoples Med | Critical Care | Discharge | Death |
| *Transition start point* | Prehospital | 0.848 | 0.084 | 0.068 |  |  |  |  |  |  |
|  | ED |  | 0.707 | 0.164 | 0.015 | 0.045 | 0.000 | 0.020 | 0.045 | 0.004 |
|  | AMU |  |  | 0.042 | 0.028 | 0.199 | 0.066 | 0.003 | 0.658 | 0.004 |
|  | Surgical |  | 0.026 |  | 0.058 | 0.081 | 0.010 | 0.027 | 0.785 | 0.012 |
|  | Speciality Med |  | 0.005 | 0.075 |  | 0.033 | 0.002 | 0.018 | 0.837 | 0.030 |
|  | Internal Med |  | 0.006 | 0.025 | 0.010 |  | 0.029 | 0.020 | 0.862 | 0.047 |
|  | Older Peoples Med |  | 0.002 | 0.011 | 0.005 | 0.042 |  | 0.002 | 0.879 | 0.059 |
|  | Critical Care |  | 0.001 | 0.343 | 0.130 | 0.334 | 0.002 |  | 0.036 | 0.154 |

AMU: Acute Medical Unit. ED: Emergency Department

**Table 3.**
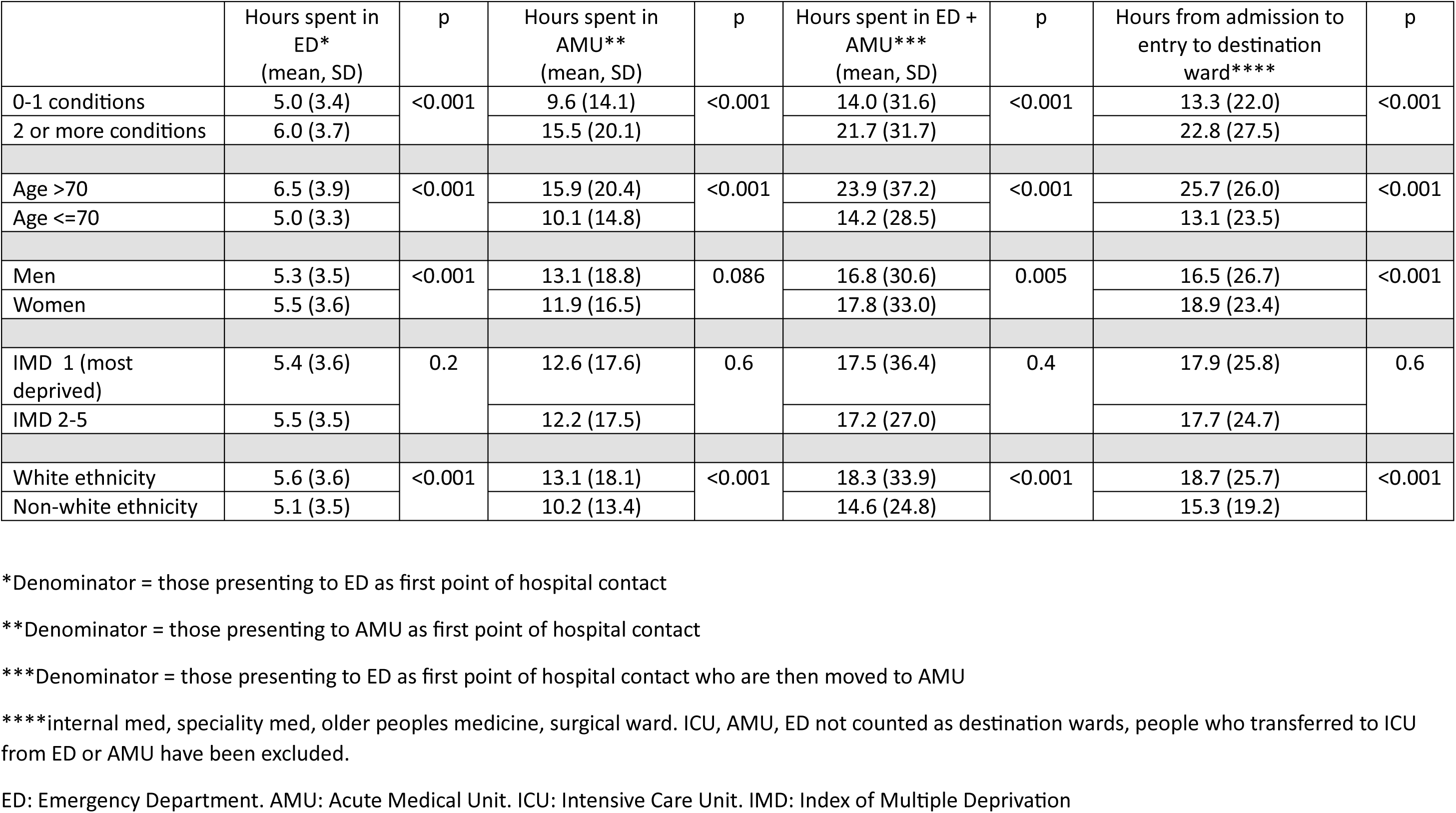
Time spent in assessment areas (emergency department and acute medical unit) prior to transfer to definitive ward of care – analysis comparing subgroups.

|  | Hours spent in ED*<br>(mean, SD) | p | Hours spent in AMU**<br>(mean, SD) | p | Hours spent in ED + AMU***<br>(mean, SD) | p | Hours from admission to entry to destination ward**** | p |
| --- | --- | --- | --- | --- | --- | --- | --- | --- |
| 0-1 conditions | 5.0 (3.4) | <0.001 | 9.6 (14.1) | <0.001 | 14.0 (31.6) | <0.001 | 13.3 (22.0) | <0.001 |
| 2 or more conditions | 6.0 (3.7) |  | 15.5 (20.1) |  | 21.7 (31.7) |  | 22.8 (27.5) |  |
| Age >70 | 6.5 (3.9) | <0.001 | 15.9 (20.4) | <0.001 | 23.9 (37.2) | <0.001 | 25.7 (26.0) | <0.001 |
| Age ≤70 | 5.0 (3.3) |  | 10.1 (14.8) |  | 14.2 (28.5) |  | 13.1 (23.5) |  |
| Men | 5.3 (3.5) | <0.001 | 13.1 (18.8) | 0.086 | 16.8 (30.6) | 0.005 | 16.5 (26.7) | <0.001 |
| Women | 5.5 (3.6) |  | 11.9 (16.5) |  | 17.8 (33.0) |  | 18.9 (23.4) |  |
| IMD 1 (most deprived) | 5.4 (3.6) | 0.2 | 12.6 (17.6) | 0.6 | 17.5 (36.4) | 0.4 | 17.9 (25.8) | 0.6 |
| IMD 2-5 | 5.5 (3.5) |  | 12.2 (17.5) |  | 17.2 (27.0) |  | 17.7 (24.7) |  |
| White ethnicity | 5.6 (3.6) | <0.001 | 13.1 (18.1) | <0.001 | 18.3 (33.9) | <0.001 | 18.7 (25.7) | <0.001 |
| Non-white ethnicity | 5.1 (3.5) |  | 10.2 (13.4) |  | 14.6 (24.8) |  | 15.3 (19.2) |  |

\*Denominator = those presenting to ED as first point of hospital contact
\*\*Denominator = those presenting to AMU as first point of hospital contact
\*\*\*Denominator = those presenting to ED as first point of hospital contact who are then moved to AMU
\*\*\*\*internal med, speciality med, older peoples medicine, surgical ward. ICU, AMU, ED not counted as destination wards, people who transferred to ICU from ED or AMU have been excluded.
ED: Emergency Department. AMU: Acute Medical Unit. ICU: Intensive Care Unit. IMD: Index of Multiple Deprivation

Supplementary Tables 4 to 8 show transition probabilities for prespecified subgroups, with both absolute and relative differences between each pair of subgroups. People with MLTC were more likely to die, be transferred to internal medicine wards or older people’s medicine wards, less likely to be transferred to surgical wards, and were less likely to be discharged directly from the acute medical unit rather than be transferred to another ward first. Similar patterns were seen for older vs younger people. Few marked differences were evident between men and women except for women being more likely to be transferred to older people’s medicine wards. No marked differences were seen between those in the bottom fifth of deprivation index and the comparator group. People of non-white ethnicity were less likely to die and were also less likely to be admitted to critical care or older people’s medicine wards.

Supplementary Table 9 shows the differences between people with and without MLTC, stratified by age group. For people aged ≤70 years, those with MLTC were more likely to die, to be transferred to internal medicine wards from AMU, and were less likely to be discharged directly home from AMU or ED than people aged ≤70 years without MLTC. These contrasts were much less evident on inspection of the matrices comparing those with and without MLTC for the subgroup of people aged over 70 years.

Table 4 shows differences in the time spent in emergency department and acute assessment areas prior to transfer to a definitive place of care for people with different characteristics. People with MLTC took longer to reach a definitive place of care and a major contributor to this was the difference in the time spent on the acute medical unit. This difference was also evident in time from hospital entry to definitive ward for those aged over 70 compared with those aged 70 or under. Smaller, yet statistically significant, differences for this metric were also observed for women vs men (longer for women) and for people of white vs non-white ethnicity (longer for white ethnicity).

Supplementary Table 10 shows the proportion of people discharged within one day of admission (i.e. those with ‘rapid turnaround’) and the proportion of people undergoing at least one move from their definitive ward to another inpatient ward during their stay (i.e. ‘boarding’). People with MLTC were less likely to return home within a day of admission than people without MLTC; a similar pattern was evident for older vs younger people. Men and those of white ethnicity were also less likely to be discharged within a day than women and those of non-white ethnicity, respectively. People with the lowest fifth of IMD scores were significantly more likely to be discharged within a day than those from higher IMD groups. Boarding was more common for people with MLTC than those without MLTC, people who were older vs younger and for men compared with women.

## Discussion

Our analysis reinforces that MLTC are common in people admitted to hospital and shows that people living with MLTC follow different pathways and have worse outcomes from hospital care than those without MLTC. People with MLTC were more likely to die in hospital, be transferred to internal medicine wards or older people’s medicine wards, were less likely to be transferred to surgical wards, and less likely to be discharged directly home from the acute medical unit. These differences were more marked for younger patients than for those aged >70 years.

Comorbidity has been studied as a modifier of care pathways and outcomes for single conditions; such as heart failure and cancer [20,21] but few studies have examined associations of MLTC with pathways of care in hospital. MLTC were associated with higher 30-day mortality, longer emergency department stays and more frequent early revisits in an analysis of Scottish healthcare data [9]. Similar to our findings, this analysis also found differences in how these associations were patterned in younger vs older groups. The close relationship between age and MLTC makes it difficult to disentangle the extent to which our findings can be attributed to the presence of MLTC as opposed to age or age-related factors e.g. frailty [22]. However, the marked difference in pathways seen for people with and without MLTC aged <70 years suggests that MLTC rather than age drives care pathway differences in this younger group.

Although it is reassuring that people with MLTC admitted to hospital do not stay much longer in the emergency department (which is known to associated with higher short-term mortality) [23,24], the longer stay in AMU before reaching a place of definitive care is still potentially of concern. Our findings align with those from a recent snapshot audit from 152 UK hospitals [25] showing that people living with frailty had delayed time to assessment and senior review, perhaps due to such patients having complex needs but lower acuity. Moves within hospital are known to be associated with worse outcomes, e.g. falls, wound infection, medication errors [26] and longer length of stay [27]. Arguably people living with MLTC are the least appropriate group to be moved in this way given their already longer length of stay, worse outcomes and lower satisfaction with care [5].

Our analysis had a number of strengths and limitations. These include use of a large dataset from one of the largest hospitals in the UK hospital serving a population with mixed ethnicity and significant socioeconomic deprivation. We had detailed, time-resolved data on ward moves and used a prespecified set of conditions and code lists to define conditions and MLTC. Our study was conducted at a single centre in a single country, and results may therefore not be generalisable to other centres, populations or health care systems, although our length of stay findings align with a recent national UK audit [25]. We confined our analysis to a single year before the COVID pandemic and it is possible that some aspects of care pathways may have changed in the post-COVID era [28].

We simplified our analysis by focussing on people admitted overnight to hospital for unscheduled care; this made analyses more tractable but gives only a partial description of the complex pathways in unscheduled hospital care as between 40% and 80% of emergency department (ED) attendances do not result in hospital admission [9]. Further analyses placing ED attendance at the centre of the analyses would add additional insights. Additional descriptors of pathways of care would add richness and detail beyond our use of wards and future analyses should try to incorporate these.

We chose to categorise variables including age and ethnicity to make analyses tractable with adequate sample sizes. This inevitably led to some loss of detail and we acknowledge variability within categories. There is no ‘gold standard’ for pathways of care in hospital for people living with MLTC and thus in contrast with our previous work [12], we can compare subgroups with each other rather than with an agreed standard of care.

Our findings have important implications for current practice and for hospital system redesign. Pathways of care followed by people with MLTC are different, and several of our findings suggest that their care processes are suboptimal. Further research is needed to understand the reasons for these variations in pathways of care from the perspective of patients, clinicians and hospital systems [7]. Future analyses could examine how pathways of care have changed in the post-COVID era, how pathways differ between hospitals, and whether people with different combinations of long-term conditions follow different pathways. Our analyses could also be extended to elective admissions and to incorporate a broader range of data (e.g. time spent in each place of care). Understanding pathways of care for people with MLTC admitted to hospital is an essential first step; the challenge now is to work with patients and clinicians to reimagine processes and pathways of care to improve outcomes and patient experience for the growing number of people admitted to hospital who are living with MLTC.

## Acknowledgements

The work of the ADMISSION Research Collaborative uses data provided by patients and collected by the NHS as part of their care and support. MDW, RC, SB and AAS acknowledge support from the National Institute for Health and Care Research (NIHR) Newcastle Biomedical Research Centre based at Newcastle upon Tyne Hospitals NHS Foundation Trust, Cumbria, Northumberland, Tyne and Wear NHS Foundation Trust and Newcastle University (ref: NIHR203309). MDW, RC, JS and AAS acknowledge support from the Multiple Long-Term Conditions Cross-NIHR Collaboration. MDW acknowledges support from the NIHR Newcastle Clinical Research Facility. SP and JS are funded by the NIHR HealthTech Research Centre in Diagnostic and Technology Evaluation. ES is supported by NIHR Midlands Patient Safety Research Collaborative and the Birmingham NIHR Biomedical Research Centre. ES, SG and FE are supported by the HDRUK PIONEER Data Hub.

## Authors Contributions

FE: Data extraction and analysis, first draft of manuscript

SB: Interpretation of results; critical revision of manuscript

RC: Conception, Funding acquisition, interpretation of results; critical revision of manuscript

SG: Data provision, Interpretation of results; critical revision of manuscript

SP: Interpretation of results; critical revision of manuscript

ES: Conception, Funding acquisition, data provision, interpretation of results; critical revision of manuscript

AAS: Conception, Funding acquisition, interpretation of results; critical revision of manuscript

JS: Interpretation of results; critical revision of manuscript

MDW: Conception, funding acquisition, interpretation of results, first draft of manuscript

## Ethics approval and consent to participate

The study was conducted under ethical approval provided for PIONEER, the Health Date Research Hub for Acute Care (East Midlands–Derby [20/EM/0158] and Confidentiality Advisory Group (20/CAG/0084). CAG approval was granted to conduct these analyses without the need for consent from included individuals.

## Availability of data

Data used in this analysis are hosted by the HDRUK Acute Data Hub (PIONEER). Data may be available on application to PIONEER subject to approval from the PIONEER data access committee and completion of appropriate data sharing agreements.

## Supplementary material

**Supplementary Table 1.** Ontology of place of care categories used in analysis.

| ward code | ward | Amended Ward | ward group | Analysis Category |
| --- | --- | --- | --- | --- |
| WAMU | Acute Medical Unit |  | Acute Medical Unit | Acute Medical Unit |
| AMSB | Ambmaj SubWait |  | ED | ED |
| AMWR | AmbMaj Waiting Room |  | ED | ED |
| ACA_A | Ambulance Decision Area |  | ED | ED |
| AmbQ | Ambulance Q |  | ED | ED |
| WACE | Ambulatory Care Episode |  | Acute Medical Unit | Acute medical unit |
| Q_Bumb_A | Ambulatory Major Bumble Bee |  | ED | ED |
| Q_InnWR_A | Ambulatory major Inner Waiting Rm |  | ED | ED |
| Q_PaedWR_A | Ambulatory Major Paeds Waiting Rm |  | ED | ED |
| Q_Psych_A | Ambulatory Major Psych Review |  | ED | ED |
| Q_Reclin_A | Ambulatory Major Recliners |  | ED | ED |
| MajAmbTria | Ambulatory Major Waiting 4 Triage |  | ED | ED |
| MajAmbWR_A | Ambulatory Major Waiting Room |  | ED | ED |
| ASKae_A | Ask A&E |  | ED | ED |
| BRVL | Bournville Ward (ERU) |  | Older People | Older People's Medicine |
| 676 | BRVL (Bournville) |  | Older People | Older People's Medicine |
| 172 | CCH (Centre for Clinical Haematology) |  | Haematology | Specialty Medicine |
| CCH | Centre Clinical Haematology |  | Haematology | Specialty Medicine |
| AmbQCold_A | Cold Ambulance Queue |  | ED | ED |
| CMajorsQ | Cold Majors Queue |  | ED | ED |
| Resus | Cold Resus |  | ED | ED |
| CVWait | Cold Waiting Room |  | ED | ED |
| GPMain | Cubicles |  | ED | ED |
| WDIS | Discharge Lounge |  | Discharge Lounge | Discharged |
| 6 | East 2A |  | Older People | Older People's Medicine |
| E2A | East 2A |  | Older People | Older People's Medicine |
| 7 | East 2B |  | Older People | Older People's Medicine |
| E2B | East 2B |  | Older People | Older People's Medicine |
| EDAA3 | EDAA |  | ED | ED |
| EDAA2 | EDAA - Subwait |  | ED | ED |
| EDAA1 | EDAA - Waiting For Triage |  | ED | ED |
| WEOU | Emergency Observations Unit |  | Acute medical unit | Acute medical unit |
| 687 | EPOC | Enhanced Perioperative Care | Critical Care | Critical Care |
| EPOC | EPOC | Enhanced Perioperative Care | Critical Care | Critical Care |
| GPWait | GP Wait Area |  | ED | ED |
| HRBN | Harborne Ward (Geri med) |  | Older People | Older People's Medicine |
| AmbQHot_A | Hot Ambulance Q |  | ED | ED |
| MajSub | Hot Major Subwait |  | ED | ED |
| Major | Hot Majors |  | ED | ED |
| HMajorQ | Hot Majors Queue |  | ED | ED |
| CMWFT | Hot Majors Waiting for Triage |  | ED | ED |
| MainWr | Hot Majors Waiting Room |  | ED | ED |
| CV2 | Hot Resus |  | ED | ED |
| 540 | HRBN (Harborne) |  | Older People | Older People's Medicine |
| Majornew | Major |  | ED | ED |
| MajAmb_A | Majors Ambulatory |  | ED | ED |
| Amb1 | Minors Waiting Room |  | ED | ED |
| NHS111_A | NHS111 |  | ED | ED |
| QHDR | QHDR | High Dose Brachytherapy |  | Speciality Medicine |
| RAT999 | RAT 999 |  | ED | ED |
| RATAMB | RAT Ambulatory |  | ED | ED |
| RATCub | RAT Cubicles |  | ED | ED |
| RATHold | RAT Holding |  | ED | ED |
| RATMain | RAT Main Wait |  | ED | ED |
| RATSub | RAT SubWait |  | ED | ED |
| Resus_A | Resus |  | ED | ED |
| 523 | Short Surge |  | Acute Medical Unit/ED | Acute Medical Unit |
| QESS | Short Surge |  | Acute Medical Unit/ED | Acute Medical Unit |
| W03 | W03 - Edgbaston |  | Older People | Older People's Medicine |
| 331 | W03 (Edgbaston) |  | Older People | Older People's Medicine |
| 967 | W302 |  | Surgery | Surgery |
| W302 | W302 |  | Surgery | Surgery |
| 962 | W303 |  | Renal | Internal Medicine |
| W303 | W303 |  | Renal | Internal Medicine |
| 961 | W304 |  | Cardiology | Internal Medicine |
| W304 | W304 |  | Cardiology | Internal Medicine |
| 958 | W305 |  | Renal | Internal Medicine |
| W305 | W305 |  | Renal | Internal Medicine |
| 953 | W306 |  | Liver | Internal Medicine |
| W306 | W306 |  | Liver | Internal Medicine |
| 951 | W407 |  | Neuro | Speciality Medicine |
| W407 | W407 |  | Neuro | Speciality Medicine |
| 949 | W408 |  | Medicine | Internal Medicine |
| W408 | W408 |  | Medicine | Internal Medicine |
| 943 | W409 |  | Neurosurgery | Surgery |
| W409 | W409 |  | Neurosurgery | Surgery |
| 942 | W410 |  | T&O | Surgery |
| W410 | W410 |  | T&O | Surgery |
| 936 | W411 |  | Neuro | Speciality Medicine |
| W411 | W411 |  | Neuro | Speciality Medicine |
| 933 | W412 |  | T&O | Surgery |
| W412 | W412 |  | T&O | Surgery |
| 931 | W513 |  | Diabetes | Internal Medicine |
| W513 | W513 |  | Diabetes | Internal Medicine |
| 926 | W514 |  | Stroke | Internal Medicine |
| W514 | W514 |  | Stroke | Internal Medicine |
| 925 | W515 |  | Respiratory | Internal Medicine |
| W515 | W515 |  | Respiratory | Internal Medicine |
| 924 | W516 |  | Respiratory | Internal Medicine |
| W516 | W516 |  | Respiratory | Internal Medicine |
| 923 | W517 |  | Medicine | Internal Medicine |
| W517 | W517 |  | Medicine | Internal Medicine |
| 918 | W518 |  | Medicine | Internal Medicine |
| W518 | W518 |  | Medicine | Internal Medicine |
| 916 | W620 |  | Surgery | Surgery |
| W620 | W620 |  | Surgery | Surgery |
| 913 | W621 |  | Surgical Assessment Unit | Surgery |
| W621 | W621 |  | Surgical Assessment Unit | Surgery |
| 907 | W622 |  | Oncology | Speciality Medicine |
| W622 | W622 |  | Oncology | Speciality Medicine |
| 902 | W623 |  | Oncology | Speciality Medicine |
| W623 | W623 |  | Oncology | Speciality Medicine |
| 173 | W623 Day Case |  | Oncology | Speciality Medicine |
| 901 | W624 |  | Haematology | Speciality Medicine |
| W624 | W624 |  | Haematology | Speciality Medicine |
| 888 | W625 |  | Haematology | Speciality Medicine |
| W625 | W625 |  | Haematology | Speciality Medicine |
| 885 | W726 |  | Mixed | Surgery |
| W726 | W726 |  | Mixed | Surgery |
| 873 | W727 |  | Mixed | Surgery |
| W727 | W727 |  | Mixed | Surgery |
| 567 | W727H |  | Mixed | Surgery |
| 727H | W727H |  | Mixed | Surgery |
| 872 | W728 |  | Mixed | Surgery |
| W728 | W728 |  | Mixed | Surgery |
| 545 | WACE |  | Acute Medical Unit | Acute Medical Unit |
| 671 | WADM | Admission Unit | Acute Medical Unit | Acute Medical Unit |
| WADM | WADM Admission Unit | Admission Unit | Acute Medical Unit | Acute Medical Unit |
| 573 | WAMB |  | Ambulatory Care | Acute Medical Unit |
| WAMB | WAMB |  | Ambulatory Care | Acute Medical Unit |
| 988 | WAMU | Acute Medical Unit | Acute Medical Unit | Acute Medical Unit |
| 908 | WAOU |  | Oncology | Speciality Medicine |
| WAOU | WAOU |  | Oncology | Speciality Medicine |
| 565 | WARC Acute Renal Care |  | Renal | Internal Medicine |
| WARC | WARC Acute Renal Care |  | Renal | Internal Medicine |
| 972 | WAST |  | Acute Medical Unit | Acute Medical Unit |
| WAST | WAST |  | Acute Medical Unit | Acute Medical Unit |
| 606 | WBU |  | Burns | Surgery |
| WBU | WBU |  | Burns | Surgery |
| 673 | QSSU |  | Short Stay Surgery | Surgery |
| QSSU | QSSU Short Stay Surgery Unit |  | Short Stay Surgery | Surgery |
| WCCA | WCCA |  | Critical Care | Critical care |
| 652 | WCCA (Critical Care A) |  | Critical Care | Critical care |
| WCCB | WCCB |  | Critical Care | Critical care |
| 654 | WCCB (Critical Care B) |  | Critical Care | Critical care |
| WCCC | WCCC |  | Critical Care | Critical care |
| 655 | WCCC (Critical Care C) |  | Critical Care | Critical care |
| WCCD | WCCD |  | Critical Care | Critical care |
| 656 | WCCD (Critical Care D) |  | Critical Care | Critical care |
| WCCE | WCCE |  | Critical Care | Critical care |
| 605 | WCCU |  | Cardiology | Internal Medicine |
| WCCU | WCCU |  | Cardiology | Internal Medicine |
| 623 | WCDU | Acute Medical Unit | Acute Medical Unit | Acute Medical Unit |
| WCDU | WCDU | Acute Medical Unit | Acute Medical Unit | Acute Medical Unit |
| 971 | WDIS | Discharge Lounge | Discharge Lounge | Discharged |
| 171 | WEOU | Emergency Observation Unit | Acute Medical Unit/ED | Acute Medical Unit |
| 571 | WSAU |  | Surgical Assessment Unit | Surgery |
| WSAU | WSAU |  | Surgical Assessment Unit | Surgery |
| 16 | WW1 |  | Older People | Older People's Medicine |
| WW1 | WW1 |  | Older People | Older People's Medicine |
| 17 | WW2 |  | Older People | Older People's Medicine |
| WW2 | WW2 |  | Older People | Older People's Medicine |
| Excluded from analysis – procedures, day case and outpatient facilities |  |  |  |  |
| 408 | WCAR |  | Cardiology daycase | Exclude |
| WCAR | WCAR |  | Cardiology daycase | Exclude |
| QCRF | Clinical Research Facility |  | Research facility | Exclude |
| QDDC | Diabetic Day Case |  | Diabetes | Exclude |
| 4 | E1A Oncology Treatment suite |  |  | Exclude |
| E1A | E1A Oncology Treatment suite |  |  | Exclude |
| 5 | E1B Infusion Suite |  | Infusion Suite | Exclude |
| E1B | E1B Infusion Suite |  | Infusion Suite | Exclude |
| EBDU | East Block Day Unit (EBDU) |  | Day Case Surgery | Exclude |
| 91 | EBDU (East Day Unit) |  | Day Case Surgery | Exclude |
| 93 | GFTS | Ground Floor Treatment Suite | Infusion Suite | Exclude |
| GFTS | Ground Floor Treatment Suite |  | Infusion Suite | Exclude |
| WIRD | Interventional Radiology |  | Interventional Radiology | Exclude |
| WPDC | Medical Tattooing Day Case-QEB |  | Plastics | Exclude |
| 987 | Norman Power |  | Step-Down (exclude) | Exclude |
| TNPC | Norman Power |  | Step-Down (exclude) | Exclude |
| QOTH | Ophthalmology Treat Suite-2 |  | Outpatient | Exclude |
| QOTS | Ophthalmology Treatment Suite |  | Outpatient | Exclude |
| WPRM | Pleural Respiratory Medicine |  |  | Exclude |
| 337 | QCRF |  | Research facility | Exclude |
| 984 | QCYT |  | Urology Daycase | Exclude |
| QCYT | QCYT |  | Urology Daycase | Exclude |
| 407 | QDDC | Diabetes Day Case |  | Exclude |
| 522 | QLTH |  | Urology Daycase | Exclude |
| QLTH | QLTH |  | Urology Daycase | Exclude |
| 674 | QMW2 |  | Urology Daycase | Exclude |
| QMW2 | QMW2 |  | Urology Daycase | Exclude |
| 170 | QOTH | Ophthalmology Treat Suite-2 | Outpatient | Exclude |
| 344 | QOTS | Ophthalmology Treatment Suite | Outpatient | Exclude |
| RINF | Renal Infusion Suite |  | Renal Infusion Suite | Exclude |
| 978 | RINF (Renal Infusion) |  | Renal Infusion Suite | Exclude |
| 952 | Virtual SAU |  | Virtual - Exclude | Exclude |
| SAUV | Virtual SAU |  | Virtual - Exclude | Exclude |
| 970 | W301 |  | Renal Dialysis | Exclude |
| W301 | W301 |  | Renal Dialysis | Exclude |
| WLI | Waiting List Initiative (BMI) |  |  | Exclude |
| 528 | WDRM |  | Dermatology daycase | Exclude |
| WDRM | WDRM |  | Dermatology daycase | Exclude |
| 568 | WEND | Endoscopy Unit | Endoscopy | Exclude |
| WEND | WEND Endoscopy Unit | Endoscopy Unit | Endoscopy | Exclude |
| 569 | WENO | Endocrine Day cases |  | Exclude |
| WENO | WENO Endocrine Day Cases | Endocrine Day cases |  | Exclude |
| 983 | WIRD | Interventional Radiology |  | Exclude |
| WLCC | WLCC (WLICC) | Waiting List Initiative CC | Exclude | Exclude |
| 406 | WPDC | Medical Tattooing Day Case-QEB | Plastics | Exclude |
| 814 | WPRM | Pleural Respiratory Medicine | Respiratory Daycase | Exclude |
| 968 | WXRY |  | Pain Management | Exclude |
| WXRY | WXRY |  | Pain Management | Exclude |
| D623 | YPU Daycase | Young Person's Unit Daycase |  | Exclude |

**Supplementary Table 2.** Descriptive data for all adults with a first admission for unscheduled care to the Queen Elizabeth Hospital between 01/07/2018 and 30/06/2019 (n=33252)

|  |  |  |  |
| --- | --- | --- | --- |
| Mean age (years) (SD) |  |  | 56.0 (21.9) |
| Age (years) (n, %) | ≤70 |  | 22920 (68.9) |
|  | >70 |  | 10332 (31.1) |
| Sex (n, %) | Women |  | 17241 (51.8) |
| Ethnicity (n, %) | White |  | 21889 (65.8) |
|  | Black |  | 1480 (4.5) |
|  | Asian |  | 5037 (15.1) |
|  | Other (including Mixed) |  | 1425 (4.3) |
|  | Unknown/ Preferred not to say |  | 3421 (10.3) |
| Index of Multiple Deprivation (n, %) | 1 (most deprived) |  | 16113 (48.5) |
|  | 2 |  | 7232 (21.7) |
|  | 3 |  | 5207 (15.7) |
|  | 4 |  | 2671 (8.0) |
|  | 5 (least deprived) |  | 1720 (5.2) |
|  | Not recorded |  | 309 (0.9) |
| Number of conditions documented prior to index admission (n, %) | 0 | No MLTC | 15438 (46.4) |
|  | 1 |  | 3796 (11.4) |
|  | 2-3 | MLTC | 5637 (17.0) |
|  | 4-5 |  | 3719 (11.2) |
|  | 6-8 |  | 3023 (9.1) |
|  | 9+ |  | 1639 (4.9) |
| Number of admissions over the last 10 years (median, IQR) |  |  | 8 (1, 12) |

**Supplementary Table 3.** Recorded prevalence of long-term conditions in the analysis population.

| Condition | Men<br>(n=16011) | Women<br>(n=17241) |
| --- | --- | --- |
| Stroke | 451 (2.8%) | 434 (2.5%) |
| Coronary artery disease | 2414 (15.1%) | 1853 (10.7%) |
| Heart failure | 918 (5.7%) | 953 (5.5%) |
| Peripheral artery disease | 473 (3.0%) | 298 (1.7%) |
| Heart valve disorders | 778 (4.9%) | 861 (5.0%) |
| Arrhythmia | 1685 (10.5%) | 1678 (9.7%) |
| Venous thromboembolic disease | 432 (2.7%) | 548 (3.2%) |
| Aneurysm | 182 (1.1%) | 78 (0.5%) |
| Hypertension | 4357 (27.2%) | 4934 (28.6%) |
| Diabetes mellitus | 2220 (13.9%) | 2228 (12.9%) |
| Addisons disease | 16 (0.1%) | 25 (0.1%) |
| Cystic fibrosis | 5 (<0.1%) | <5* (<0.1%) |
| Thyroid disorders | 391 (2.4%) | 1457 (8.5%) |
| Chronic obstructive pulmonary disease | 1170 (7.3%) | 1313 (7.6%) |
| Asthma | 1372 (8.6%) | 2447 (14.2%) |
| Bronchiectasis | 183 (1.1%) | 236 (1.4%) |
| Obstructive sleep apnoea | 356 (2.2%) | 222 (1.3%) |
| Parkinson's syndromes | 142 (0.9%) | 91 (0.5%) |
| Epilepsy | 474 (3.0%) | 465 (2.7%) |
| Multiple sclerosis | 59 (0.4%) | 98 (0.6%) |
| Paralysis | 62 (0.4%) | 44 (0.3%) |
| Peripheral neuropathy | 374 (2.3%) | 414 (2.4%) |
| Chronic primary pain | 26 (0.2%) | 52 (0.3%) |
| Solid organ cancers | 1309 (8.2%) | 1165 (6.8%) |
| Haematological cancers | 125 (0.8%) | 100 (0.6%) |
| Metastatic cancers | 216 (1.3%) | 224 (1.3%) |
| Melanoma | 43 (0.3%) | 32 (0.2%) |
| Dementia | 458 (2.9%) | 739 (4.3%) |
| Schizophrenia | 278 (1.7%) | 212 (1.2%) |
| Depression | 1121 (7.0%) | 1930 (11.2%) |
| Anxiety | 684 (4.3%) | 1195 (6.9%) |
| Bipolar disorder | 121 (0.8%) | 210 (1.2%) |
| Drug alcohol misuse | 1094 (6.8%) | 457 (2.7%) |
| Eating disorder | 6 (0.0%) | 51 (0.3%) |
| Autism | 57 (0.4%) | 37 (0.2%) |
| Post-traumatic stress disorder | 39 (0.2%) | 30 (0.2%) |
| Connective tissue disease | 297 (1.9%) | 718 (4.2%) |
| Osteoarthritis | 933 (5.8%) | 1899 (11.0%) |
| Long term musculoskeletal problems due to injury | 6 (<0.1%) | 6 (<0.1%) |
| Osteoporosis | 160 (1.0%) | 777 (4.5%) |
| Gout | 458 (2.9%) | 179 (1.0%) |
| Chronic liver disease | 545 (3.4%) | 425 (2.5%) |
| Inflammatory bowel disease | 195 (1.2%) | 244 (1.4%) |
| Chronic pancreatic disease | 128 (0.8%) | 52 (0.3%) |
| Peptic ulcer | 337 (2.1%) | 295 (1.7%) |
| Diverticular disease | 849 (5.3%) | 1246 (7.2%) |
| Gastro oesophageal reflux disease | 938 (5.9%) | 1205 (7.0%) |
| Chronic kidney disease | 1095 (6.8%) | 1287 (7.5%) |
| End stage kidney disease | 300 (1.9%) | 240 (1.4%) |
| Hyperplasia of the prostate | 1030 (6.4%) | 0 (0.0%) |
| Endometriosis | 0 (0.0%) | 90 (0.5%) |
| Recurrent urinary tract infection | 197 (1.2%) | 279 (1.6%) |
| Anaemia | 1262 (7.9%) | 1875 (10.9%) |
| Uncorrectable visual impairment | 139 (0.9%) | 193 (1.1%) |
| Uncorrectable hearing impairment | 296 (1.8%) | 369 (2.1%) |
| Menieres disease | 21 (0.1%) | 45 (0.3%) |
| HIV AIDS** | - | - |
| Chronic Lyme Disease | 0 (0.0%) | 0 (0.0%) |
| Tuberculosis | 64 (0.4%) | 50 (0.3%) |
| Congenital disease and chromosomal abnormalities | 234 (1.5%) | 288 (1.7%) |

*\*True number suppressed under terms of access for PIONEER data*
*\*\*Data not available under terms of access for PIONEER data*

**Supplementary Table 4:**
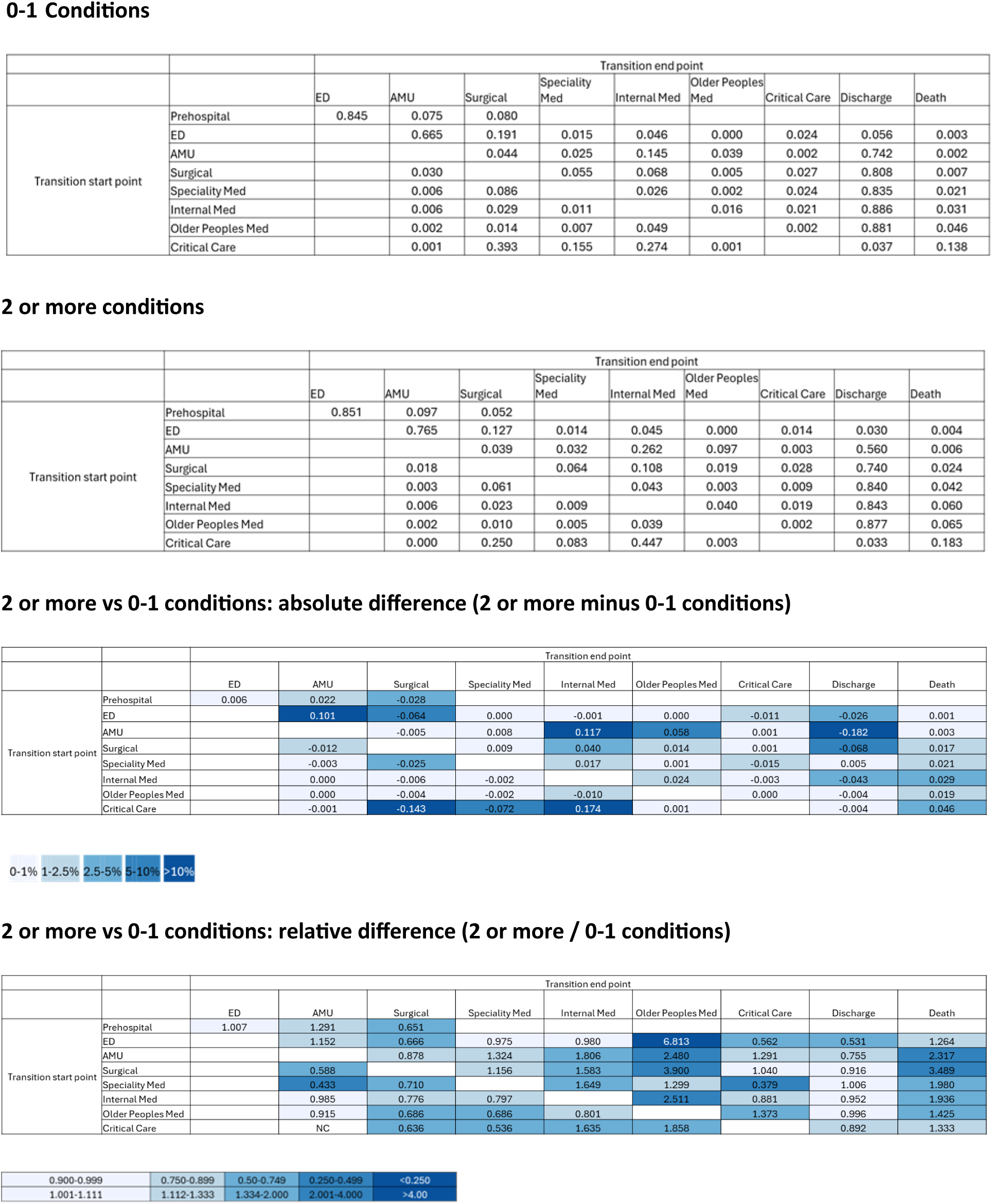
Contrasts in transition matrices for people with multiple long-term conditions (2 or more conditions) vs those with 0-1 long-term conditions.

**Supplementary Table 5:**
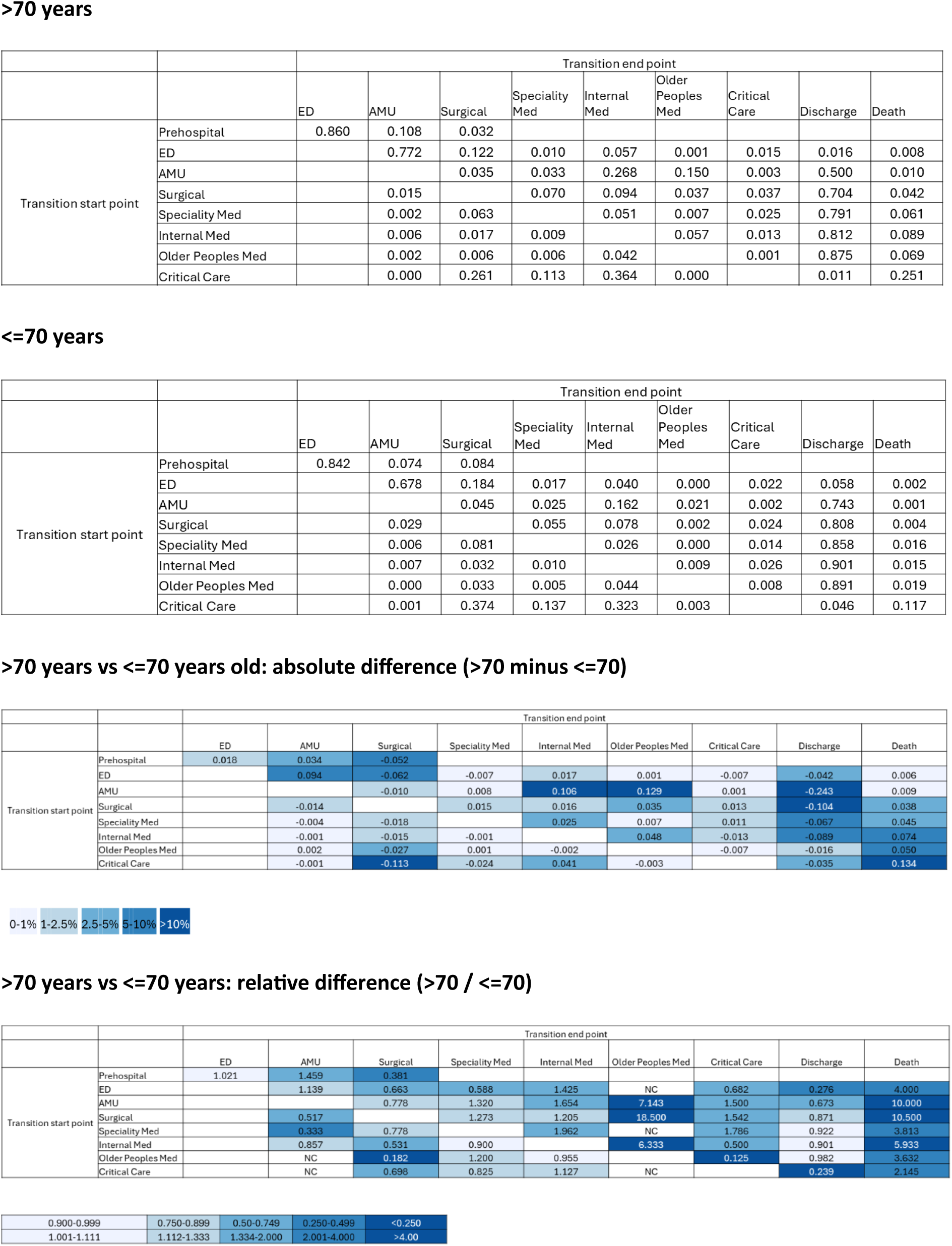
Contrasts in transition matrices for people aged >70 years vs <=70 years.

**Supplementary Table 6:**
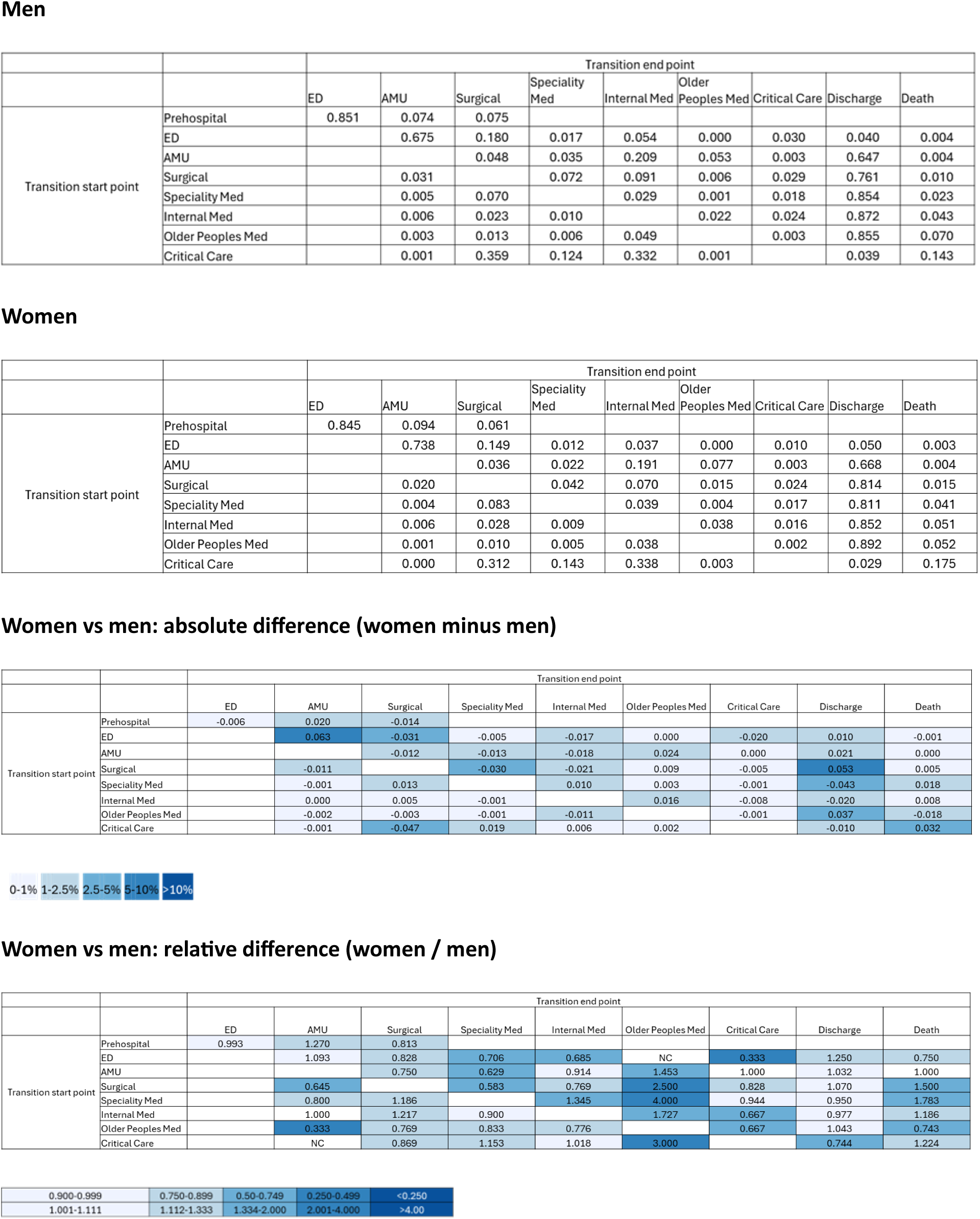
Contrasts in transition matrices for men vs women Men.

**Supplementary Table 7:**
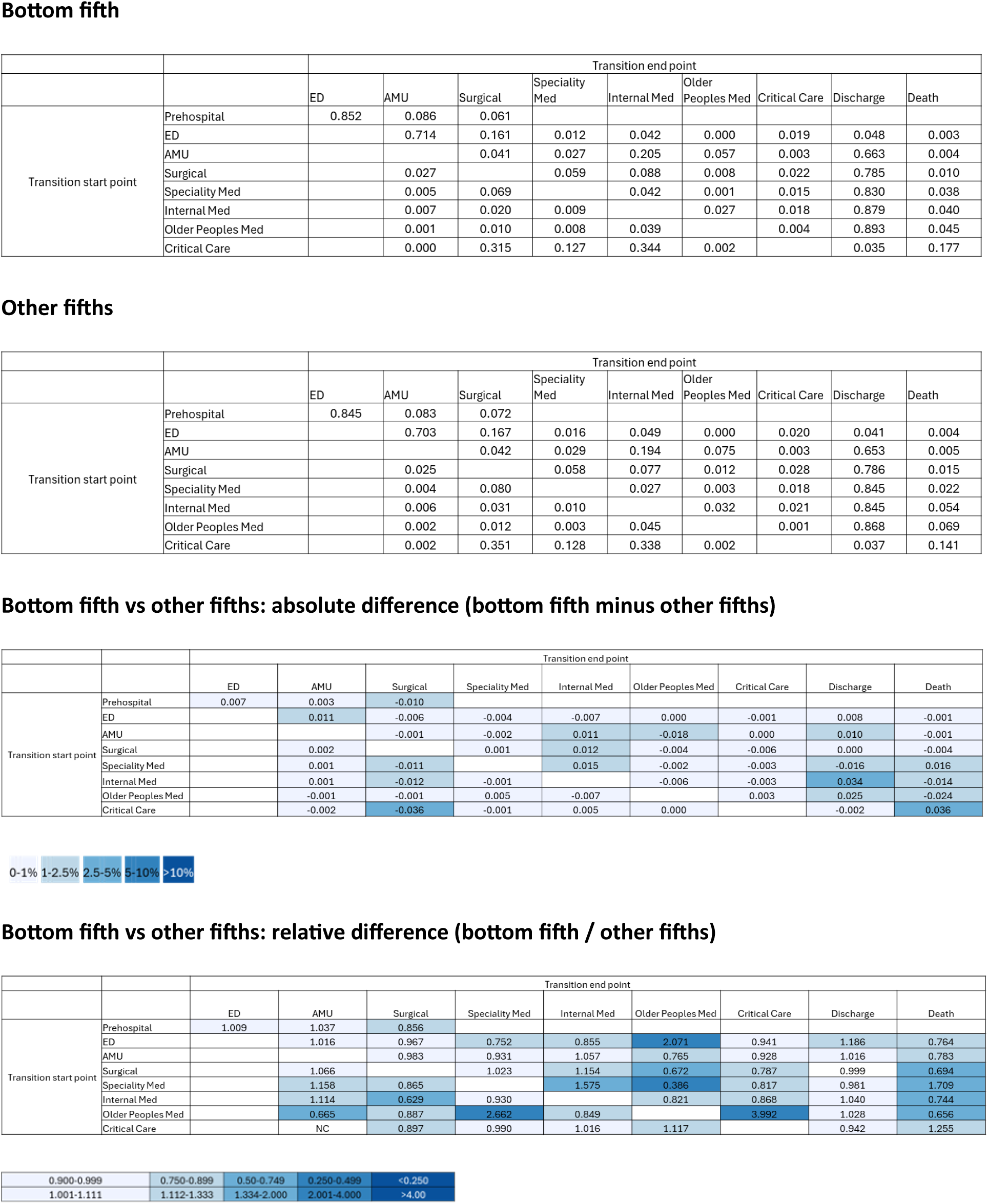
Contrasts in transition matrices for people in the bottom fifth of UK deprivation vs people in the other four fifths.

**Supplementary Table 8:**
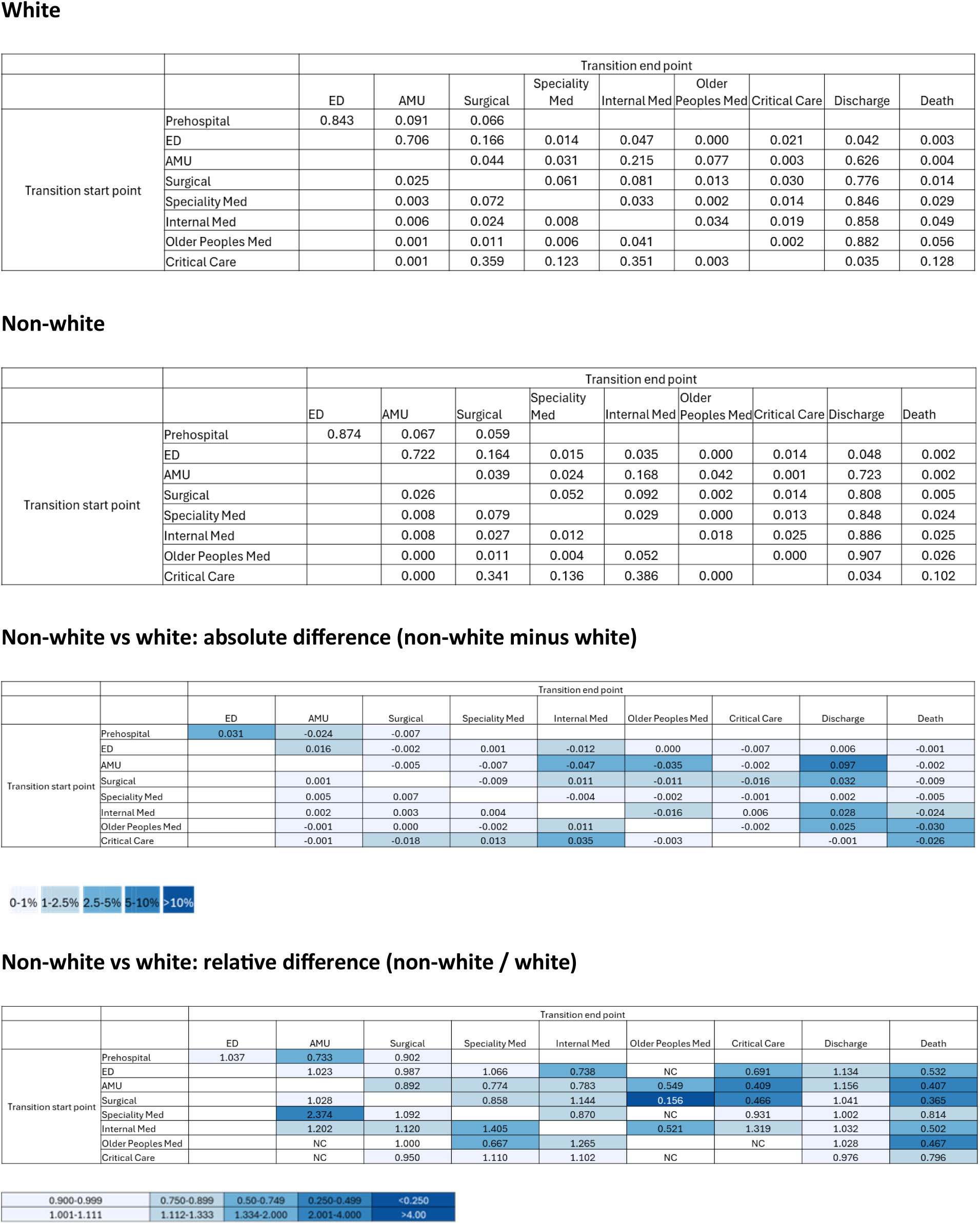
Contrasts in transition matrices for people of white vs non-white ethnicity.

**Supplementary Table 9:**
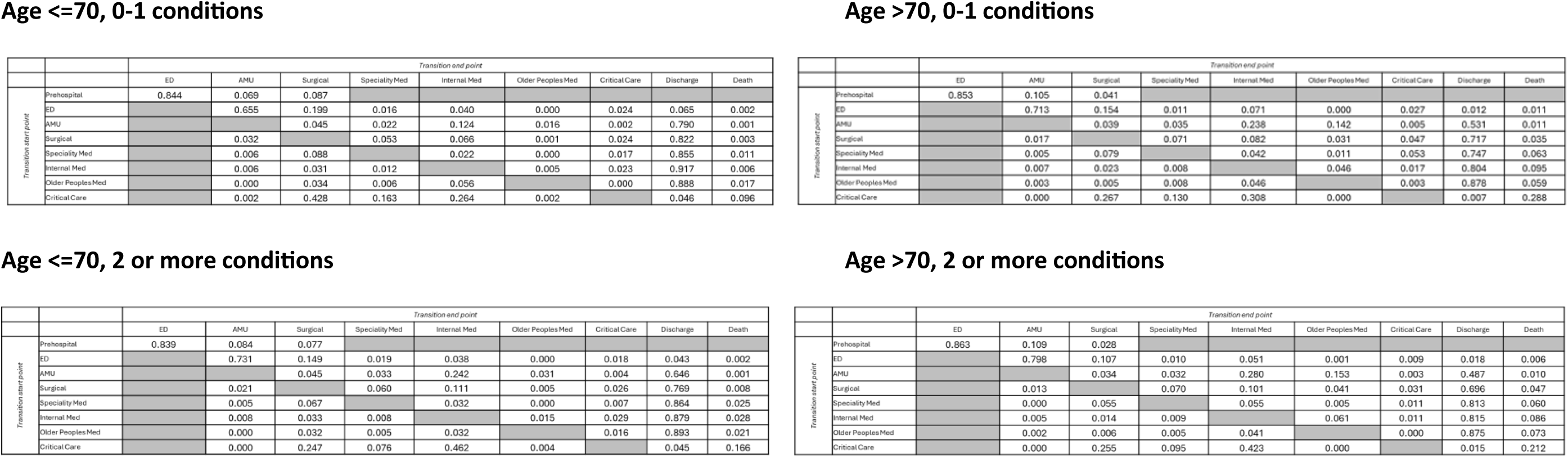
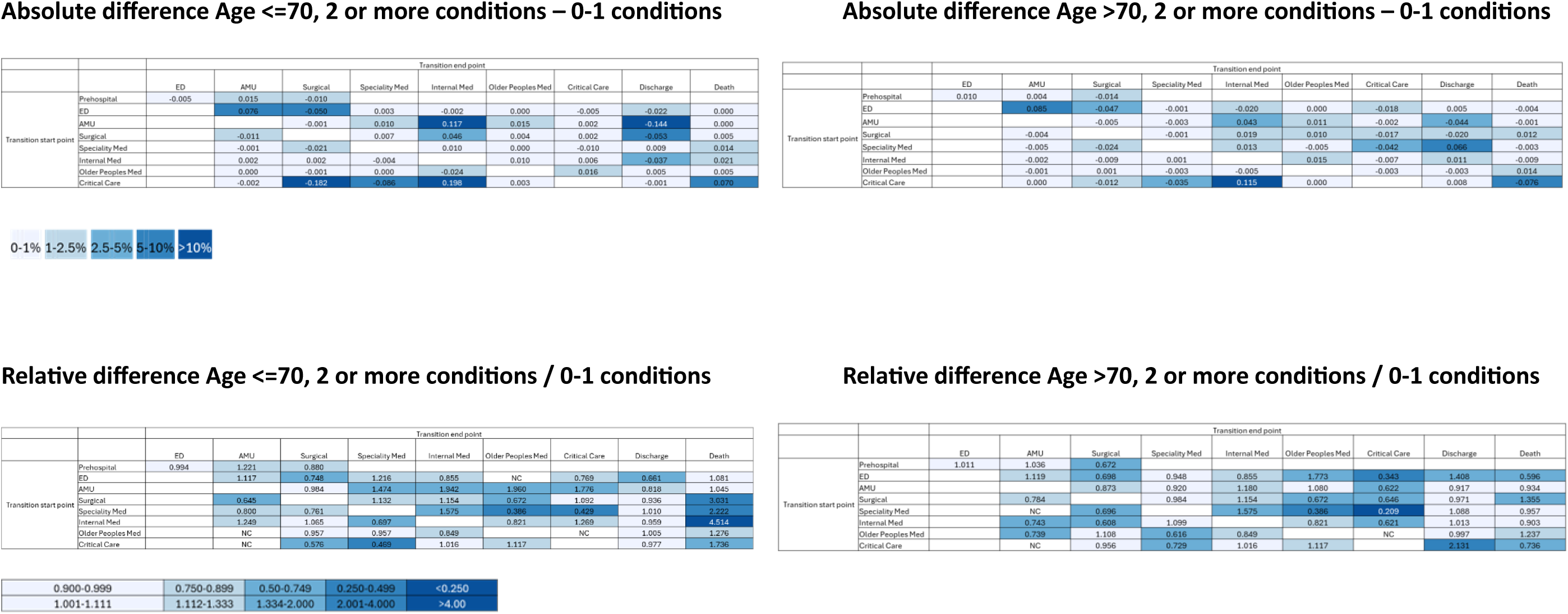
Contrasts in transition matrices for people with 0-1 long-term conditions vs those with multiple long-term conditions (2 or more conditions), stratified by age group.

**Supplementary Table 10.** Proportion of people with rapid discharge (defined as a hospital stay of 1 day or less) and with moves from definitive place of care to a different ward (boarding) – analysis comparing subgroups.

|  | Proportion with total hospital LOS of 1 day or less (%) | p | Proportion moved from a ward to another ward* | p |
| --- | --- | --- | --- | --- |
| 0-1 conditions | 10934 (56.8%) | <0.001 | 1489 (16.4) | 0.002 |
| 2 or more conditions | 5583 (39.8%) |  | 1389 (18.2) |  |
| Age >70 | 3374 (32.7%) | <0.001 | 1126 (18.7) | <0.001 |
| Age ≤70 | 13143 (57.3%) |  | 1752 (16.4) |  |
| Men | 7612 (47.5%) | <0.001 | 1609 (18.7) | <0.001 |
| Women | 8904 (51.6%) |  | 1269 (15.6) |  |
| IMD 1 (most deprived) | 8171 (50.7%) | <0.001 | 1323 (16.8) | 0.13 |
| IMD 2-5 | 8194 (48.7%) |  | 1529 (17.7) |  |
| White ethnicity | 10094 (46.1%) | <0.001 | 2036 (17.7) | 0.20 |
| Non-white ethnicity | 4606 (58.0%) |  | 599 (16.8) |  |

IMD: Index of Multiple Deprivation. LOS: Length of stay
\*Denominator is all those admitted to the following categories of ward: internal med, speciality med, older peoples med, surgical ward
Ward to ward move = a move to another ward in these four categories. Moves to ICU, AMU, ED, discharge or death are not counted. Moves within a category (e.g. from one internal med ward to another internal med ward) are counted as moves

**Supplementary Figure 1.**
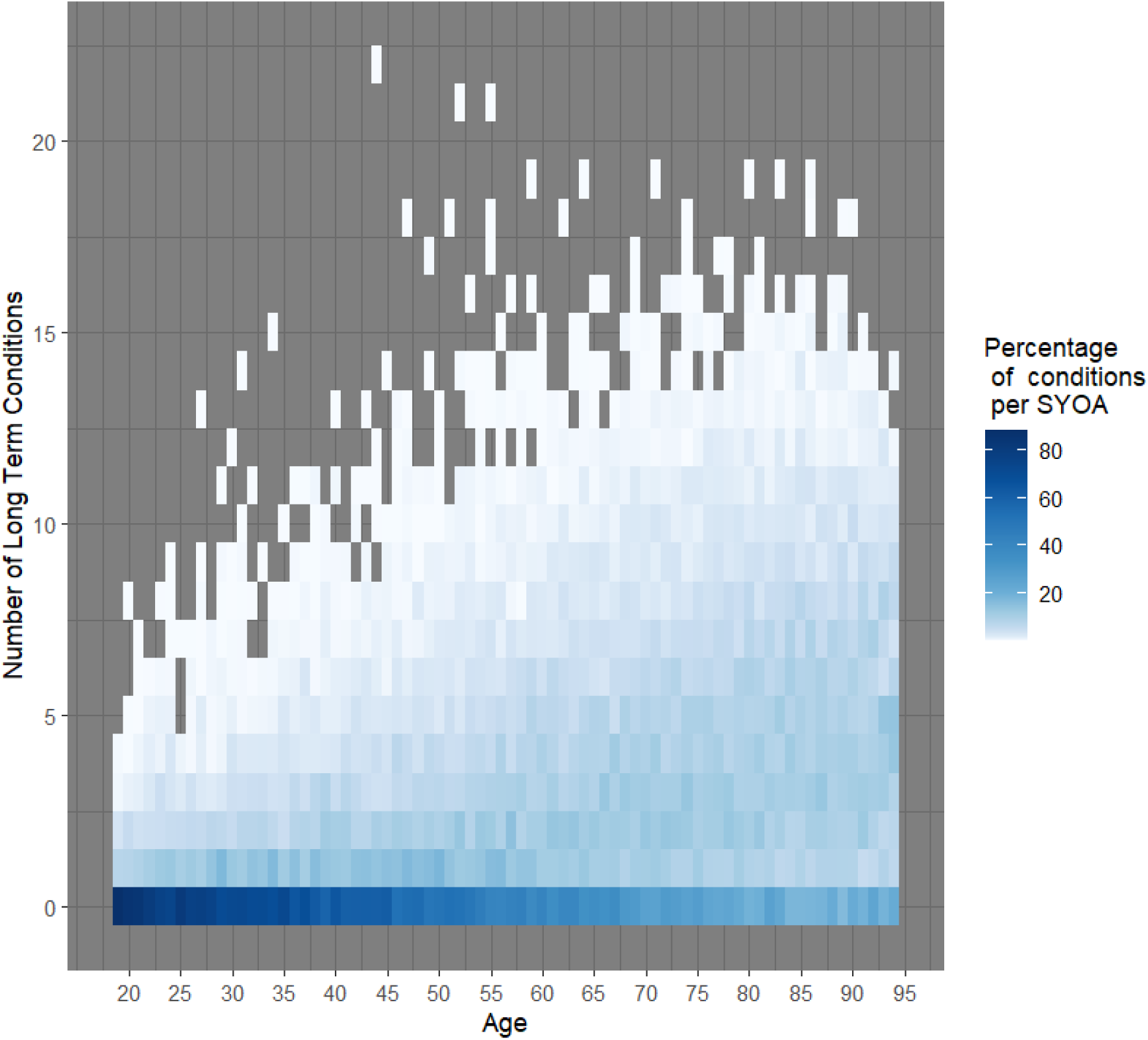
Association between age and number of long-term conditions in the analysis cohort. Spearmans’ Rho = 0.493 SYOA: Single Year of age

**Supplementary Figure 2:**
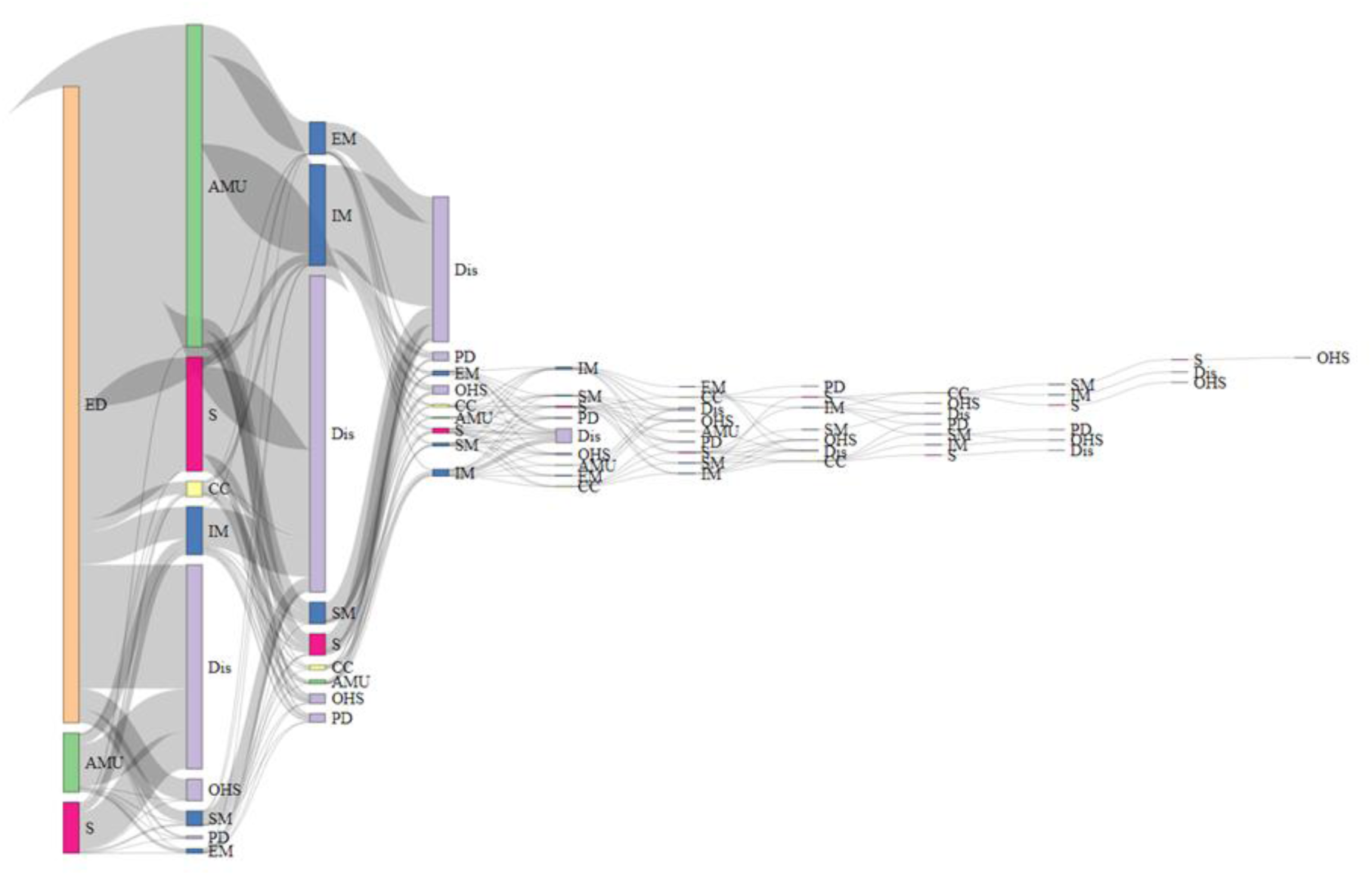
Sankey diagram depicting ward moves through hospital admission. Sankey diagram for unscheduled care admissions where the first place of care was the emergency department, acute medical unit or surgical ward. ED = Emergency Department, AMU = Acute Medical Unit, S= Surgery, CC = Critical Care, IM = Internal Medicine, SM = Speciality Medicine, EM = Older Person Care, Dis = Discharge, OHS = Other Hospital Service (e.g. transfer to another hospital, step-down wards, virtual wards), PD = Person Died

